# Associated factors of acculturation strategies and mental health outcomes among international students in China

**DOI:** 10.1101/2024.09.08.24313277

**Authors:** Chenchang Xiao, Jingyan Yan, Hanjia Li, Changmian Ding, Bin Yu

## Abstract

**Objectives:** There is an increasing number of international students in China. Acculturation strategies are the way students cope with different cultures, including integration, assimilation, separation, and marginalization. This study aims to investigate the acculturation strategies and associated factors, and the effect of these strategies on mental health status among international students in China.

**Study design:** Cross-sectional study.

**Methods:** Study data were collected from 567 international students attending universities in China. Acculturation strategies, acculturative stress, depressive symptoms were measured using reliable and valid scales. Linear and multinomial logistic regression were used for analysis.

**Results:** Study findings revealed that integration (31.57%) was the most preferred acculturation strategy, followed by marginalization (28.92%), separation (21.87%) and assimilation (17.54%). Females were more likely to choose separation strategy than marginalization, while students with religions had higher likelihood to choose integration strategy. Students majoring in Literature/Art and liking their major were more likely to use assimilation strategy. Students with more studying time in weekdays and medium studying time in weekends were more likely to prefer integration strategy. Students with separation and integration strategy had higher acculturative stress.

**Conclusions:** Integration is the most popular acculturation strategy among international students in China. Students with separation and integration strategy had worse mental health status. Gender, major, religion, daily study time were significantly associated with the preference of acculturation strategies.

## Introduction

### International students in China

With the development of economic globalization, China has attracted a large and increasing number of international students to study and work. By the end of 2017, there was a total of 489,200 international students from 204 countries studying in China^32^. Studying abroad provides international students with opportunities to access quality education and expand intercultural competence^3^. However, adapting to a new cultural, social and educational environment is challenging and stressful. International students may encounter a lot of challenges, including language barriers, loneliness, homesickness, and academic failures^4–6^. These difficulties may increase the risk of developing mental health problems, such as acculturative stress and depressive symptoms.

### Acculturation and acculturation strategies

Acculturation is a challenging process of change in values, beliefs, attitudes, and behaviors when individuals interact with different cultures^7, 8^. Acculturation strategies are the different coping styles taken by individuals when they live with or contact with two cultures^9^. Acculturation strategies are developed based on Berry’s bi-dimensional acculturation: one is the identification with the heritage culture, and another is the identification with the host culture. Thus, four acculturation strategies are generated, including integration, assimilation, separation, and marginalization (Fig 1)^7^. Integration refers to the strategy that individuals maintain the heritage culture and adopt the host culture simultaneously, while the marginalization strategy comes true for adopting neither of heritage nor host cultures. Assimilation is the strategy that people adopt the host culture but reject the heritage culture, while separation is maintaining the heritage culture but rejecting the host culture. Different acculturation strategies may affect the process of acculturation for international students^7, 10, 11^.

**Fig 1.**
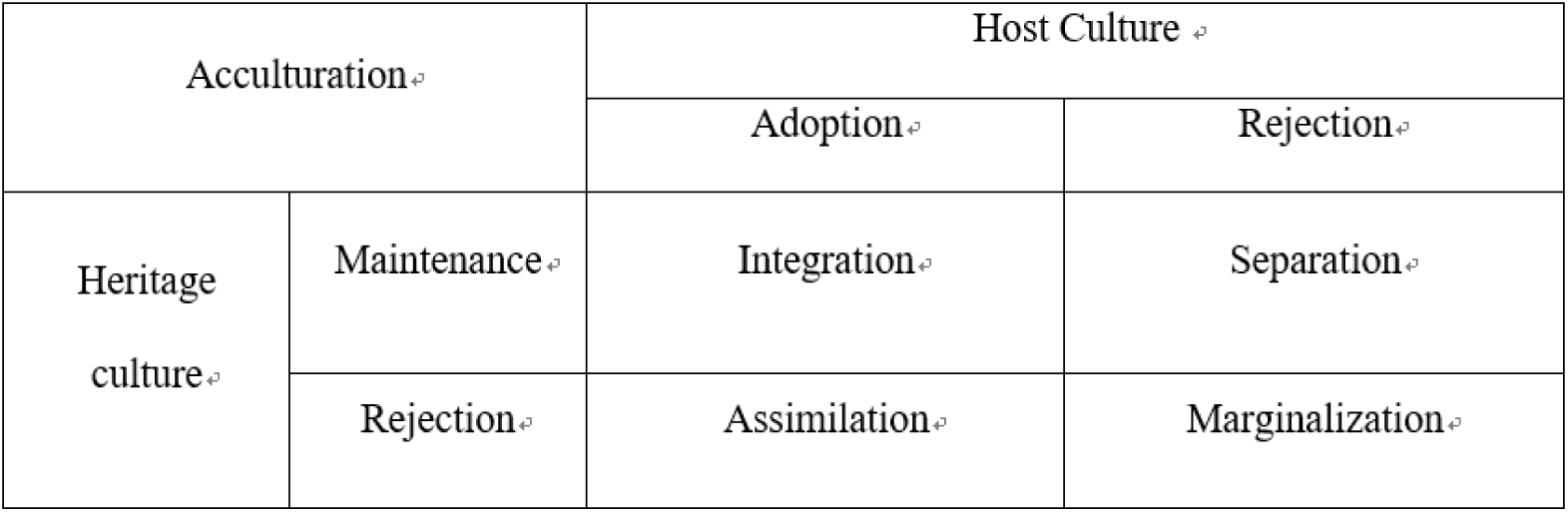
Acculturation strategies based on the attitudes towards the host culture and heritage culture

### Effect of acculturation strategies on mental health status

Acculturation strategies have been found to be significantly associated with mental health outcomes, especially acculturative stress and depression^20–22^. For example, one study conducted among 200 North Indian migrated students in Bangalore city of Karnataka found that students with separation and marginalization strategies had higher acculturative stress^23^. Another study among Turkish immigrants in Germany showed that people with integration strategy had the lowest level of depression, while people with marginalization had the highest^24^. International students, as a special mobile population, experienced high levels of acculturative stress and depression^27–30^. However, to our knowledge, few studies have explored the relationships between acculturation strategies with mental health problems among international students in China. Therefore, one aim of this study is to investigate the effects of different acculturation strategies on mental health status.

### Associated factors of acculturation strategies

An overwhelming empirical studies in acculturation have focused on the effects of different acculturation strategies on various outcomes^13, 14^. However, limited studies have investigated the associated factors of acculturation strategies^14, 15^. One study conducted among 210 Turkish migrants showed that individuals who were young, female and with higher education often applied a participatory strategy (e.g. integration and assimilation), while those aged 50 or older and less educated often applied non-participating strategy (e.g. separation and marginalization)^16^. Another study conducted among 313 immigrant adolescents in Portugal showed that ethnicity, majority identity, discrimination, experience adaptation, gender, age were associated with acculturation strategy preference^12^. Other studies also reported that international students who were married and lived with their spouses may experience sociocultural segregation because a substantial part of their time was invested in being with their spouses^17, 18^. Another study conducted among international students in China found that male and longer residence significantly predicted lower degree of acculturation^19^. However, few studies have investigated the effect of study-related factors (e.g., major, study time, etc.) on the preference of acculturation strategies. The main purpose of the study is to investigate the demographic and study-related factors of acculturation strategies among international students in China.

### Purpose of the study

The present study aims to (1) investigate the preference of acculturation strategies among international students in China; (2) examine the associated factors of acculturation strategies; (3) analyze the associations between acculturation strategies with acculturative stress and depression. The ultimate goal is to provide data for future effective interventions on mental health among international students in China.

## Methods

### Participants and sampling

Participants in the study were international students who were attending universities in Wuhan, China with foreign nationality. Wuhan is a university city with 82 colleges and universities with over 19,000 international students were studying in Wuhan in 2016^33^. Four largest universities in Wuhan, covering nearly 60% of the total international students, were selected to recruit participants^34^.

Detailed sampling processes were described elsewhere^5, 34^. Briefly, firstly, four universities were selected from Wuhan, including Wuhan University, Huazhong University of Science and Technology, Central China Normal University, and Zhongnan University of Economics and Law. Secondly, in each selected university, classes of international students, the smallest unit for schooling in China, were randomly selected. All international students in China were grouped into classes to study Chinese mandarin with approximately 40-50 students per class. All international students in the selected classes were eligible to participate. The study was voluntary, and only the students who signed the written informed consent were recruited. A total of 630 international students were recruited and completed the survey.

### Data collection

Data were collected through the self-developed International Student Health and Behavior Survey (ISHBS). The ISHBS is a packet of questionnaires that asked about demographic and study-related information, acculturative stress, acculturation strategies, and depression. Both English and Chinese versions of the questionnaire were available for participants to choose from. No significant difference was found between the data collected using Chinese version (n=62) and English version (n=505)^5^.

The survey was conducted in September 1st in 2012 and ended in November 30th. The survey was implemented in a classroom with a vacant seat placed between two students to ensure personal privacy and independence. At the end of survey, all participants were asked to put their questionnaire into an envelope and placed it into a pre-prepared locked box. Data were manually entered into the computer with Epidata software. Double-entry protocol was used for minimizing data-entry errors. Among 630 international students, 63 were excluded due to the missing data for several key variables, yielding 567 (90%) participants in the final sample. The study was approved by the institutional review board at Wuhan University, China.

### Measures

#### Acculturation Strategy

Acculturation strategy was measured using the self-developed Acculturation Strategy Assessment (ASA) (supplementary Table S1). ASA was developed based on Berry’s four strategies of acculturation^7, 35^, and several developed scales measuring acculturation strategies^7, 36–38^. ASA consists of 30 items assessing two dimensions of acculturation with 15 items for each dimension. One dimension measures the host culture adaptation, another dimension measures the self-culture maintenance. A seven-point Likert scale was used to measure the ASA ranging from 1 (strongly disagree) to 7 (strongly agree). Total score for each dimension was computed. Median score for each dimension was used as cut-off point. Participants scored high on both dimensions were categorized as “Integration”, and participants who scored low on both were categorized as “Marginalization”. Participants were grouped into “Assimilation” if they scored high on host culture subscale, and low on self-culture subscale, and participants were grouped into “Separation” if scored opposite. The Cronbach alpha was 0.87 for total ASA, and 0.86 for host culture subscale and 0.85 for self-culture subscale.

#### Acculturative Stress

Acculturative stress was measured using the Acculturative Stress Scale for International Students (ASSIS)^39^. ASSIS is a widely used scale with 36 items, measured using a five-point Likert scale ranging from 1 (strongly disagree) to 5 (strongly agree). In our previous study^5^, we categorized the ASSIS into seven subscales, including value conflict (VC), identity threat (IT), opportunity deprivation (OD), mistreated (MT), lack of cultural competence (CC), lack of self-confidence(CS), and homesickness (HS) based on the item content, item response, exploratory and confirmative factor analysis. Total score was estimated with higher score indicating greater level of acculturative stress. The Cronbach alpha was 0.93.

#### Depression

Depression was measured using the 10-item Centers for Epidemiological Studies Depression Scale (CESD-10)^40^. A four-point Likert scale was used to assess individual items with 0 (rarely or none of the time, or less than1 day) to 3 (most of the time or 5–7 days). Two items were reversely coded, and total scores were computed with higher scores indicating greater depression. The Cronbach alpha was 0.70.

##### Demographic variables and covariates

Demographic variables and other covariates in the study included age (in years), gender (male/female), place of origin (Asia/Africa/others), marital status (unmarried/married/divorced /separated/others; and was categorized into unmarried/others), education (undergraduate /graduate), religion (Christian/Muslim/Hinduism or Buddhism/Judaism/ no religion/others), duration in China (in months; and was categorized into one year or less/more than one year), major area (sciences/language and art/medicine/business/law/others), if like the major (don’t like/don’t know/like), level of preparedness before coming to China (not well/well), number of roommates (0/1/2/3 or more), study time in weekdays/weekends (0-2 hours/3-4 hours/5-6 hours/> 6 hours).

### Statistical analysis

Descriptive analyses (e.g., frequency, mean, standard deviation) were used to describe the sample characteristics. Chi-square analysis was used to compare the frequency of four acculturation strategies across different covariates. Multinomial logistic regression was used to investigate the associated factors of four acculturation strategies with marginalization as reference group. The covariates with p value less than 0.1 in the Chi-square test were included in the multinomial logistic regression analysis. Linear regression was used to investigate the associations between acculturation strategies with acculturative stress and depression when controlling for covariates. All statistical analyses were conducted using SAS version 9.4 (SAS Institute, Cary, NC, USA).

## Results

### Sample characteristics

Among the total 567 participants, 336 (59.26%) were males with mean age of 22.75 (SD=4.11) years (Table 1). 84.24% of participants came from Africa or Asia, and 89.30% were unmarried. 76.32% of participants were undergraduate students, and the average time of living in China was 14.45 months (SD=12.11). Christian (35.77%) and Muslim (32.92%) were the most common religious beliefs. Literature/Art and Medicine were the top two majors with 80% of the students liked their major, and 72.69% were well prepared. 24.12% were living alone, and half and one third of them were studying 5 or more hours during weekdays and weekends, respectively.

**Table 1.** Characteristics of the study sample

| **Variables** | **Male** | **Female** | **Total** |
| --- | --- | --- | --- |
| **Total, n (%)** | 336 (59.26) | 231 (47.04) | 567 (100.00) |
| **Age, n (%)** |  |  |  |
| 20 or less | 99 (29.46) | 100 (43.29) | 199 (35.10) |
| 21-25 | 160 (47.62) | 106 (45.89) | 266 (46.91) |
| >25 | 77 (22.92) | 25 (10.82) | 102 (17.99) |
| Mean (SD) | 23.36 (4.43) | 21.85 (3.40) | 22.75 (4.11) |
| **Place of origin, n (%)** |  |  |  |
| Asia | 132 (40.24) | 110 (49.11) | 242 (43.84) |
| Africa | 147 (44.82) | 76 (33.93) | 223 (40.40) |
| Others | 49 (14.94) | 38 (16.96) | 87 (15.76) |
| **Marital status, n (%)** |  |  |  |
| Unmarried | 291 (87.65) | 210 (91.70) | 501 (89.30) |
| Others | 41 (12.35) | 19 (8.30) | 60 (10.70) |
| **Education, n (%)** |  |  |  |
| Undergraduate | 240 (74.30) | 179 (79.20) | 419 (76.32) |
| Graduate | 83 (25.70) | 47 (20.80) | 130 (23.68) |
| **Religion, n (%)** |  |  |  |
| Christian | 113 (33.73) | 88 (38.77) | 201 (35.77) |
| Muslim | 133 (33.73) | 52 (22.91) | 185 (32.92) |
| Buddhist/Hindu | 43 (12.84) | 58 (25.55) | 101 (17.97) |
| Others | 4 (1.19) | 2 (0.88) | 6 (1.07) |
| None | 42 (12.54) | 27 (11.89) | 69 (12.28) |
| **Duration in China, n (%)** |  |  |  |
| One year or less | 239 (71.13) | 171 (74.03) | 410 (72.31) |
| More than one year | 97 (28.87) | 60 (25.97) | 157 (27.69) |
| Mean (SD), months | 14.77 (12.28) | 13.98 (11.88) | 14.45 (12.11) |
| **Major, n (%)** |  |  |  |
| Science/Engineering | 57 (12.27) | 21 (9.21) | 78 (13.98) |
| Literature/Art | 116 (35.15) | 79 (34.64) | 195 (34.95) |
| Medicine | 101 (30.61) | 96 (42.11) | 197 (35.30) |
| Others | 56 (16.97) | 32 (14.04) | 88 (15.77) |
| **If like major, n (%)** |  |  |  |
| Don't like | 36 (11.04) | 26 (11.45) | 62 (11.21) |
| Don't know | 27 (8.28) | 21 (9.25) | 48 (8.68) |
| Like | 263 (80.67) | 180 (79.30) | 443 (80.11) |
| **Preparedness, n (%)** |  |  |  |
| Not well | 90 (29.70) | 52 (23.96) | 142 (27.31) |
| Well | 213 (70.30) | 165 (76.04) | 378 (72.69) |
| **Number of roommates, n** |  |  |  |
| 0 | 77 (23.69) | 53 (24.77) | 130 (24.12) |
| 1 | 105 (32.31) | 63 (29.44) | 168 (31.17) |
| 2 | 65 (20.00) | 35 (16.36) | 100 (18.55) |
| 3 or more | 78 (24.00) | 63 (29.44) | 141 (26.16) |
| **Study time in weekday (hours), n (%)** |  |  |  |
| 0-2 | 67 (20.30) | 51 (22.37) | 118 (21.15) |
| 3-4 | 88 (26.67) | 62 (27.19) | 150 (26.88) |
| 5-6 | 67 (20.30) | 49 (21.49) | 116 (20.79) |
| >6 | 108 (32.73) | 66 (28.95) | 174 (31.18) |
| **Study time in weekend (hours), n (%)** |  |  |  |
| 0-2 | 108 (32.53) | 80 (35.09) | 188 (33.57) |
| 3-4 | 95 (28.61) | 73 (32.02) | 168 (30.00) |
| 5-6 | 58 (17.47) | 39 (17.11) | 97 (17.32) |
| >6 | 71 (21.39) | 36 (15.79) | 107 (19.11) |

### Acculturation strategies

Table 2 shows that the proportion of marginalization, integration, assimilation and separation strategy were 28.92%, 31.57%, 17.54%, and 21.87%, respectively. Gender (χ^2^=21.61, p<0.001), religion (P<0.001*), major (χ^2^=38.94, p<0.001), if like the major (χ^2^=13.23, p=0.04), preparedness for studying in China (χ^2^=8.21, p=0.04), number of roommates (χ^2^=18.78, p=0.03), daily studying time in weekday (χ^2^=25.74, p=0.002) were significantly associated with the acculturation strategies.

**Table 2.** Associated factors of acculturation strategies among international students in China.

| **Variables** | **Marginalization** | **Separation** | **Assimilation** | **Integration** | **$\chi^2$ (p value)** |
| --- | --- | --- | --- | --- | --- |
| **Total, n (%)** | 164 (28.92) | 124 (21.87) | 100 (17.54) | 179 (31.57) |  |
| **Gender, n (%)** | | | | | $\chi^2=21.61$ (p<0.001) |
| Male | 103 (62.80) | 52 (41.94) | 70 (70.00) | 111 (62.01) |  |
| Female | 61 (37.20) | 72 (58.06) | 30 (30.00) | 68 (37.99) |  |
| **Age, n (%)** | | | | | $\chi^2=10.83$ (p=0.09) |
| 20 or less | 59 (35.98) | 54 (43.55) | 28 (28.00) | 58 (32.40) |  |
| 21-25 | 75 (45.73) | 54 (43.55) | 46 (46.00) | 91 (50.84) |  |
| >25 | 30 (18.29) | 16 (12.90) | 26 (26.00) | 30 (16.76) |  |
| **Place of origin, n (%)** | | | | | $\chi^2=8.14$ (p=0.23) |
| Asia | 61 (38.85) | 63 (52.50) | 42 (42.42) | 76 (43.18) |  |
| Africa | 67 (42.68) | 43 (35.83) | 37 (37.37) | 76 (43.18) |  |
| Others | 29 (18.47) | 14 (11.67) | 20 (20.20) | 24 (13.64) |  |
| **Marital status, n (%)** | | | | | $\chi^2=1.77$ (p=0.62) |
| Unmarried | 149 (91.98) | 108 (87.80) | 88 (88.89) | 156 (88.14) |  |
| Others | 13 (8.02) | 15 (12.20) | 11 (11.11) | 21 (11.86) |  |
| **Education, n (%)** | | | | | $\chi^2=3.08$ (p=0.38) |
| Undergraduate | 119 (74.38) | 95 (78.51) | 69 (71.13) | 136 (79.53) |  |
| Graduate | 41 (25.63) | 26 (21.49) | 28 (28.87) | 35 (20.47) |  |
| **Religion, n (%)** |  |  |  |  | P<0.001* |
| Christian | 55 (33.95) | 46 (37.10) | 35 (35.35) | 65 (36.72) |  |
| Muslim | 51 (31.48) | 39 (31.45) | 24 (24.24) | 71 (40.11) |  |
| Buddhist/Hindu | 25 (15.43) | 25 (20.16) | 21 (21.21) | 30 (16.95) |  |
| Others | 1 (0.62) | 1 (0.81) | 3 (3.03) | 1 (0.56) |  |
| None | 30 (18.52) | 13 (10.48) | 16 (16.16) | 10 (5.65) |  |
| **Duration in China, n (%)** | | | | | $\chi^2=3.65$ (p=0.30) |
| 1 year or less | 121 (73.78) | 82 (66.13) | 77 (77.00) | 130 (72.63) |  |
| More than 1 year | 43 (26.22) | 42 (33.87) | 23 (23.00) | 49 (27.37) |  |
| **Major, n (%)** | | | | | $\chi^2=38.94$ (p<0.001) |
| Science/Engineering | 25 (15.63) | 14 (11.48) | 12 (12.24) | 27 (15.17) |  |
| Literature/Art | 65 (40.63) | 25 (20.49) | 47 (47.96) | 58 (32.58) |  |
| Medicine | 49 (30.63) | 68 (55.74) | 22 (22.45) | 58 (32.58) |  |
| Others | 21 (13.13) | 15 (12.30) | 17 (17.35) | 35 (19.66) |  |
| **If like major, n (%)** | | | | | $\chi^2=13.23$ (p=0.04) |
| Don't like | 24 (15.29) | 14 (11.38) | 6 (6.06) | 18 (10.34) |  |
| Don't know | 21 (13.38) | 10 (8.13) | 7 (7.07) | 10 (5.75) |  |
| Like | 112 (71.34) | 99 (80.49) | 86 (86.87) | 146 (83.91) |  |
| **Preparedness, n (%)** | | | | | $\chi^2=8.21$ (p=0.04) |
| Not well | 52 (35.86) | 31 (26.96) | 22 (23.66) | 37 (22.16) |  |
| Well | 93 (64.14) | 84 (73.04) | 71 (76.34) | 130 (77.84) |  |
| **Number of roommate, n (%)** | | | | | $\chi^2=18.78$ (p=0.03) |
| 0 | 38 (24.68) | 21 (17.36) | 29 (30.53) | 42 (24.85) |  |
| 1 | 48 (31.17) | 31 (25.62) | 26 (27.37) | 63 (37.28) |  |
| 2 | 28 (18.18) | 22 (18.18) | 20 (21.05) | 30 (17.75) |  |
| 3 or more | 40 (25.97) | 47 (38.84) | 20 (21.05) | 34 (20.12) |  |
| **Study time in weekday (hours), n (%)** | | | | | $\chi^2=25.74$ (p=0.002) |
| 0-2 | 48 (30.19) | 24 (19.51) | 25 (25.00) | 21 (11.93) |  |
| 3-4 | 39 (24.53) | 39 (31.71) | 20 (20.00) | 52 (29.55) |  |
| 5-6 | 30 (18.87) | 19 (15.45) | 28 (28.00) | 39 (22.16) |  |
| >6 | 42 (26.42) | 41 (33.33) | 27 (27.00) | 64 (36.36) |  |
| **Study time in weekend (hours), n (%)** | | | | | $\chi^2=15.30$ (p=0.08) |
| 0-2 | 67 (41.88) | 36 (29.27) | 37 (37.00) | 48 (27.12) |  |
| 3-4 | 38 (23.75) | 34 (27.64) | 32 (32.00) | 64 (36.16) |  |
| 5-6 | 23 (14.38) | 28 (22.76) | 16 (16.00) | 30 (16.95) |  |
| >6 | 32 (20.00) | 25 (20.33) | 15 (15.00) | 35 (19.77) |  |

Note: \* fisher test;

### Associated factors of acculturation strategies

Results in Table 3 show that compared with males, female students were more likely to choose separation strategy *(OR=*1.86, *95% CI =* [1.03, 3.35]). Students with Christian (*OR =*2.61, *95% CI =* [1.01, 6.79]), Muslim (*OR =*4.55, *95% CI =* [1.65, 12.53]), and Buddhist/Hindu (*OR =*3.57, *95% CI =* [1.20, 10.65]) had higher likelihood to choose integration strategy. Students majoring in Literature/Art (*OR=*3.00, *95% CI =* [1.05, 8.59]) were more likely to use assimilation strategy, while students who like their own major preferred to utilize assimilation strategy. Integration strategy was often applied by the students whose daily studying time were 3-4 hours (*OR=*2.83, *95% CI =* [1.28, 6.30]), 5-6 hours (*OR=*3.65, *95% CI =* [1.47, 9.08]), >6 hours (*OR=*3.99, *95% CI =* [1.70, 9.39]) in weekday and 3-4 hours (*OR=*1.94, *95% CI =* [1.00, 3.79]) in weekend.

**Table 3.**
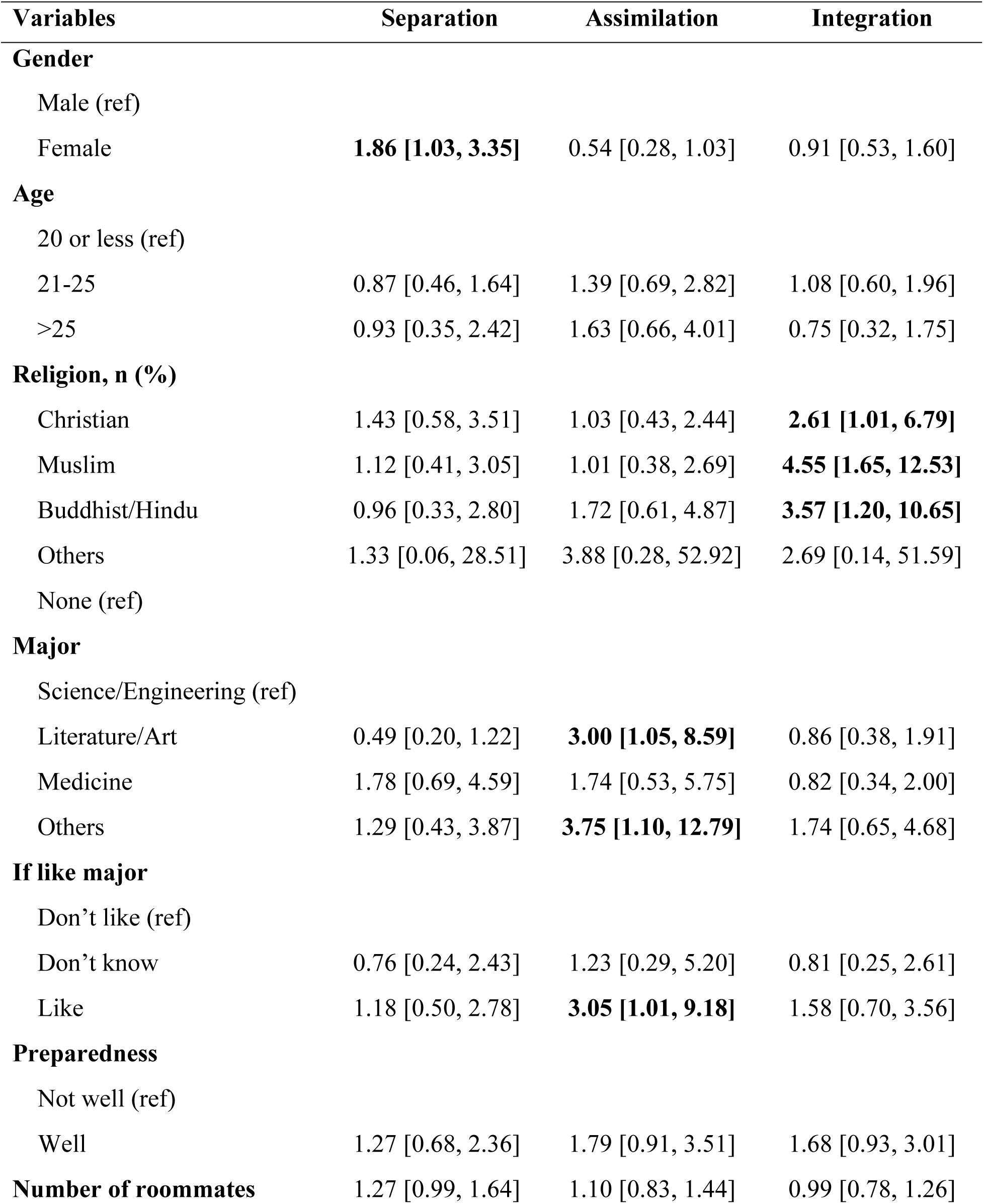
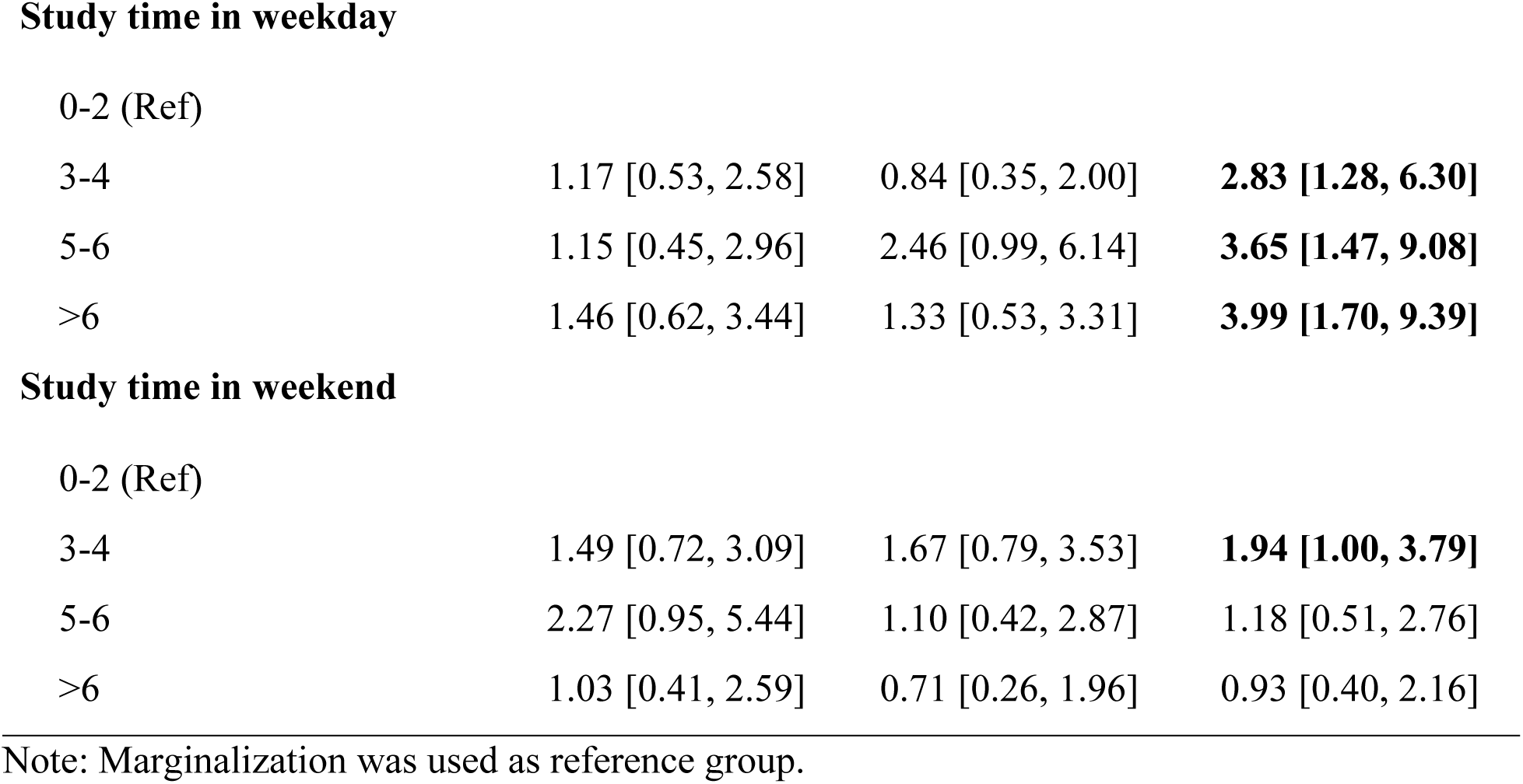
Multinomial logistic regression of acculturation strategies and associated factors, Odds Ratio [95% CI].

| **Variables** | **Separation** | **Assimilation** | **Integration** |
| --- | --- | --- | --- |
| **Gender** |  |  |  |
| Male (ref) |  |  |  |
| Female | **1.86 [1.03, 3.35]** | 0.54 [0.28, 1.03] | 0.91 [0.53, 1.60] |
| **Age** |  |  |  |
| 20 or less (ref) |  |  |  |
| 21-25 | 0.87 [0.46, 1.64] | 1.39 [0.69, 2.82] | 1.08 [0.60, 1.96] |
| >25 | 0.93 [0.35, 2.42] | 1.63 [0.66, 4.01] | 0.75 [0.32, 1.75] |
| **Religion, n (%)** |  |  |  |
| Christian | 1.43 [0.58, 3.51] | 1.03 [0.43, 2.44] | **2.61 [1.01, 6.79]** |
| Muslim | 1.12 [0.41, 3.05] | 1.01 [0.38, 2.69] | **4.55 [1.65, 12.53]** |
| Buddhist/Hindu | 0.96 [0.33, 2.80] | 1.72 [0.61, 4.87] | **3.57 [1.20, 10.65]** |
| Others | 1.33 [0.06, 28.51] | 3.88 [0.28, 52.92] | 2.69 [0.14, 51.59] |
| None (ref) |  |  |  |
| **Major** |  |  |  |
| Science/Engineering (ref) |  |  |  |
| Literature/Art | 0.49 [0.20, 1.22] | **3.00 [1.05, 8.59]** | 0.86 [0.38, 1.91] |
| Medicine | 1.78 [0.69, 4.59] | 1.74 [0.53, 5.75] | 0.82 [0.34, 2.00] |
| Others | 1.29 [0.43, 3.87] | **3.75 [1.10, 12.79]** | 1.74 [0.65, 4.68] |
| **If like major** |  |  |  |
| Don't like (ref) |  |  |  |
| Don't know | 0.76 [0.24, 2.43] | 1.23 [0.29, 5.20] | 0.81 [0.25, 2.61] |
| Like | 1.18 [0.50, 2.78] | **3.05 [1.01, 9.18]** | 1.58 [0.70, 3.56] |
| **Preparedness** |  |  |  |
| Not well (ref) |  |  |  |
| Well | 1.27 [0.68, 2.36] | 1.79 [0.91, 3.51] | 1.68 [0.93, 3.01] |
| **Number of roommates** | 1.27 [0.99, 1.64] | 1.10 [0.83, 1.44] | 0.99 [0.78, 1.26] |
| **Study time in weekday** |  |  |  |
| 0-2 (Ref) |  |  |  |
| 3-4 | 1.17 [0.53, 2.58] | 0.84 [0.35, 2.00] | **2.83 [1.28, 6.30]** |
| 5-6 | 1.15 [0.45, 2.96] | 2.46 [0.99, 6.14] | **3.65 [1.47, 9.08]** |
| >6 | 1.46 [0.62, 3.44] | 1.33 [0.53, 3.31] | **3.99 [1.70, 9.39]** |
| **Study time in weekend** |  |  |  |
| 0-2 (Ref) |  |  |  |
| 3-4 | 1.49 [0.72, 3.09] | 1.67 [0.79, 3.53] | **1.94 [1.00, 3.79]** |
| 5-6 | 2.27 [0.95, 5.44] | 1.10 [0.42, 2.87] | 1.18 [0.51, 2.76] |
| >6 | 1.03 [0.41, 2.59] | 0.71 [0.26, 1.96] | 0.93 [0.40, 2.16] |

Note: Marginalization was used as reference group.

### Effects of acculturation strategies on acculturative stress and depression

Results in Table 4 show that compared with international students with marginalization strategy, those with separation (β=11.08, p<0.001) and integration (β=6.66, p<0.05) had higher acculturative stress. Individuals with separation strategy were more likely to have higher stress in homesickness (β=3.14, p<0.001), value conflicts (β=1.77, p<0.001), mistreat (β=1.18, p<0.05), lack of cultural competence (β=1.75, p<0.001) and lack of self-confidence (β=1.34, p<0.05), while students with assimilation had higher value conflicts than marginalization (β=1.08, p<0.05), and students with integration had higher homesickness (β=1.69, p<0.001), value conflicts (β=1.40, p<0.001) and lack of cultural competence (β=1.04, p<0.05).

**Table 4.** Associations between acculturation strategies with mental health status among international students in China, regression coefficients (β)

| **Variables** | **Marginalization** | **Separation** | **Assimilation** | **Integration** |
| --- | --- | --- | --- | --- |
| ***Mental health status*** |  |  |  |  |
| **Acculturative stress** |  |  |  |  |
| Total | Ref | 11.08** | 3.30 | 6.66* |
| HS | Ref | 3.14** | 0.61 | 1.69** |
| VC | Ref | 1.77** | 1.08* | 1.40** |
| IT | Ref | 0.88 | 0.15 | 0.27 |
| OD | Ref | 0.16 | 0.20 | 0.31 |
| MT | Ref | 1.18* | 0.41 | 0.80 |
| CC | Ref | 1.75** | -0.29 | 1.04* |
| SC | Ref | 1.34* | 0.91 | 0.71 |
| **Depression** | Ref | 0.09 | -0.08 | -0.07 |

Note: <sup>1</sup>Age, gender, religion, major and study time were controlled. <sup>2</sup>HS = Homesickness, VC= Value conflict, IT= Identity threat, OD = Opportunity deprivation, MT = Mistreated, CC = Lack of cultural competence, SC = Lack of self-confidence. <sup>3</sup>\* p<0.05, \*\*p<0.001

## Discussion

This study applied Berry’s acculturation framework^7, 9^ to examine acculturation strategies of international students in China. Four acculturation strategies, including marginalization, separation, assimilation and integration, were measured. The study investigated the associated factors of the four strategies and the associations between the four strategies and mental health status among international students. Findings of the study provided information to understand the acculturation strategies, and provide evidence in developing future effective interventions on mental health among international students.

### Acculturation strategies among international students in China

Our findings reveal that integration was the most popular strategy among international students, consistent with previous studies in different countries, including China^42^, Japan^43^, Belgium^15^ and the United States^13^. Multicultural society has been gradually forming and played an important role in choice of integration strategy^15, 44, 45^. Assimilation was the least preferred strategy and less than one third of participants abandoned their heritage culture and actively participated in Chinese culture. One possible explanation was that the development of cultural values is subtle, and it would be difficult to modify or abandon specific cultures^46^. Another possible reason was that most international students prefer to return to their home countries after completing their short-term studies in China. Thus, they are less likely to change their values^15^. Marginalization and separation were found to be the moderate preferred strategies in our study, which were in line with the previous studies^43, 47^.

### Acculturation strategies and associated factors

The study investigated the associated factors of the four acculturation strategies, and identified six significant factors, including gender, religion, major, preference of major, daily studying time in weekday and weekend. Relative to males, females were more likely to choose separation strategy. Females tend to build stronger co-national ties, whose primary social network was the friendships with people from the same home country^15^. More domestic cultural contacts than international exchanges may lead to higher likelihood of adopting separation strategy.

International students majoring in Literature/Art, liking their own major were more likely to prefer the strategy of assimilation. Students majoring in Literature/Art had higher aesthetic taste, imaginative thinking, and historical logic, and were more knowledgeable in history, culture, politics, society, etc. The skills and knowledge might play important roles in cross-cultural contacts^48^. In addition, compared with students majoring in Science, Liberal Arts students had higher intercultural competence, including more intercultural communication, more confidence in dealing with situational diversities, and more likelihood to show active attitude toward various cultures^49^. Students majoring in Literature/Art were also often characterized with higher cultural awareness. They studied Chinese language and cultural arts so as to comprehensively understand Chinese culture. Therefore, they will actively participate in the culture of China and enjoy the fun during their limited study time.

Students with religions of Christian, Muslim, and Buddhist/Hindu were more likely to prefer integration strategy than marginalization. China is a multi-religious country with five major religions, including Buddhism, Taoism, Islam, Catholicism and Christianity^50^. There are more than 100 million people with religions, more than 85,000 places for religious activities, and over 3,000 religious groups in China^50^. Laws in China give people the freedom and respect of religious beliefs. International students with various religions could maintain their religion while simultaneously enjoy the Chinese culture.

Findings of the study also show that students with more time on study in weekdays were more likely to be integrated in both cultures. The students spending more time on study in weekday may actively participate in the curriculum practice and maintain interactions with teachers and classmates to improve their academic achievements. Meanwhile, these students may gather more social support through the social networks developed in the process of learning and interpersonal communication with Chinese students and local people^51^. Furthermore, it was also beneficial in improving Chinese language and deepening the understanding of Chinese culture^52^.

### Effects of acculturation strategies on mental health status

Findings of the study indicate that relative to marginalization strategy, international students with separation strategy had higher level of acculturative stress, particularly higher in homesickness, value conflict, mistreated, lack of cultural competence and lack of self-confidence. It was consistent with previous studies that individuals who separate themselves from the host culture were more likely to experience a higher level of cultural stress^35^.

Individuals who applied separation strategy tend to perceive greater cultural distance^53^, which in turn may lead to higher acculturative stress and more homesickness^54, 55^. On the other hand, findings of the study show that students with integration strategy also had higher level of acculturative stress and its components, including homesickness, value conflicts and lack of cultural competence. The results were contrary to previous studies among immigrants that people with integration strategy had the lowest acculturative stress^4, 7^. The underlying mechanism of the different results would be complicated. One possible reason was that international students, unlike immigrants, arrived in the host country for a short period of time. Previous studies have reported that individuals often experience higher acculturative stress in the first two years of immigration^56, 57^. In this study, 72.31% of international students have lived in China for only one year or less. During the process of integration, poor language communication and insufficient cultural integration may trigger different aspects of cultural conflicts, especially value conflicts. International students often feel homesickness after leaving home^58^, however, homelessness may also become stressful when the lack of cultural competence and value conflict happened^5, 39^.

### Limitations

There were limitations in the study. Firstly, it was a cross-sectional study. Longitudinal studies are needed in the future to examine the causal relationships of international students’ acculturation strategies with mental health. Secondly, the study was conducted with international students in Wuhan City. Cautions are needed when generalizing the results to the international students of the whole nation. Thirdly, data of this study were self-reported. Recall bias and social undesirability could not be ruled out.

### Conclusions

Integration is the most popular acculturation strategy among international students in China. Students with separation and integration strategy had worse mental health status. Gender, major, religion, daily study time were significantly associated with the preference of acculturation strategies. Finding in the study deepened our understanding of acculturation strategy among international students in China, and provided evidence for developing future effective interventions to prevent the mental health in this special population.

## Data Availability

The data can be available by requeat from researchers.

## Acknowledgments

The study conception and design were thought by Chenchang Xiao and Bin Yu. Data collection was conducted by Chenchang Xiao, Jingyan Yan, Hanjia Li, Changmian Ding and Bin Yu. Data analysis was conducted by Chenchang Xiao and Jingyan Yan. Drafting of manuscript was conducted by Chenchang Xiao, Jingyan Yan, Hanjia Li and Changmian Ding. Review of the manuscript was conducted by Bin Yu.

